# Assessing Vulnerability to COVID-19 in High-Risk Populations: The Role of SARS-CoV-2 Spike-Targeted Serology

**DOI:** 10.1101/2022.08.22.22279079

**Authors:** Harvey W Kaufman, William A Meyer, Nigel J Clarke, Jeff Radcliff, Christopher M Rank, James Freeman, Marcia Eisenberg, Laura Gillim, William G Morice, David M Briscoe, David S Perlin, Jay G Wohlgemuth

**Author notes:** **Correspondence:** Dr. Jay G Wohlgemuth, Chief Medical Officer and Senior VP R&D, Medical and Population Health, Quest Diagnostics, 33608 Ortega Highway, San Juan Capistrano, CA 92675.

## Abstract

**Importance:** Individuals at increased risk for severe outcomes from COVID-19, due to compromised immunity or other risk factors, would benefit from objective measures of vulnerability to infection based on prior infection and/or vaccination. We reviewed published data to identify a specific role and interpretation of SARS-CoV-2 spike-targeted serology testing for such individuals. We also provide real-world evidence of spike-targeted antibody test results, identifying the seronegativity rate across the United States from March 2021 through June 2022. Analysis of antibody test results were compared between post-transplant (ie, immunocompromised) and all other patients tested in the first half of 2022. Finally, specific recommendations are provided for an evidence-based and clinically useful interpretation of spike-targeted serology to identify vulnerability to infection and potential subsequent adverse outcomes.

**Observations:** Decreased vaccine effectiveness among immunocompromised individuals is linked to correspondingly high rates of breakthrough infections. Evidence indicates that negative results on SARS-CoV-2 antibody tests are associated with increased risk for subsequent infection. Results from widely available, laboratory-based tests do not provide a direct measure of protection but appear to correlate well with the presence of surrogate pseudovirus-neutralizing antibodies. The results of SARS-CoV-2 semiquantitative tests have also been associated with vaccine effectiveness and the likelihood of breakthrough infection. The data suggest that “low-positive” results on semiquantitative SARS-CoV-2 spike-targeted antibody tests may help identify persons at increased relative risk for breakthrough infection leading to adverse outcomes. In an analysis of data from large national laboratories during the COVID-19 Omicron-related surge in 2022, results from SARS-CoV-2 spike-targeted antibody tests were negative in 16.6% (742/4459) of solid organ transplant recipients tested compared to only 11.0% (47,552/432,481) of the remaining tested population.

**Conclusions and Relevance:** Standardized semiquantitative and quantitative SARS-CoV-2 spike-targeted antibody tests may provide objective information on risk of SARS-CoV-2 infection and associated adverse outcomes. This holds especially for high-risk populations, including transplant recipients, who demonstrate a relatively higher rate of seronegativity. The widespread availability of such tests presents an opportunity to refine risk assessment for individuals with suboptimal SARS-CoV-2 antibody levels and to promote effective interventions. Interim federal guidance would support physicians and patients while additional investigations are pursued.

## Introduction

The United States has transitioned to a new phase of the coronavirus disease-2019 (COVID-19) pandemic, with society largely reopened and vaccines and therapeutics widely available. However, due to the emergence of highly transmissible severe acute respiratory syndrome coronavirus-2 (SARS-CoV-2) Omicron variants, vaccine hesitancy/fatigue, and waning immunity in the population, transmission of SARS-CoV-2 continues. SARS-CoV-2 antibody responses to vaccination and/or infection wane over time,^1^ and there is no widely recognized, objective measure of an individual’s residual protection, including among individuals at higher than population risk.^2^ High-risk individuals are not uncommon and include those with immunocompromised conditions. Singston and colleagues estimated that immunocompromised individuals account for 12% of hospitalized COVID-19 patients.^3^ Further, immunocompromised patients had higher adjusted odds ratios for adverse outcomes, regardless of vaccination.^3^

Objective laboratory measures of SARS-CoV-2 infection risk may be clinically important for people at elevated risk for severe COVID-19 owing to compromised immunity or other risk factors.^4^ Results from laboratory-based, SARS-CoV-2 spike-targeted antibody tests, especially those quantified using standardized metrics and correlated to neutralizing antibodies (NtAb), hold potential as a critical tool for identifying individuals at the greatest infection risk. Currently, guidance for SARS-CoV-2 serology use is limited by uncertainty regarding how the results should inform clinical decision-making.^5-7^

In this Special Communication, we (1) review published data on risk factors for severe COVID-19 illness and clinical interpretation of SARS-CoV-2 spike-targeted antibody data; (2) evaluate seronegativity based on real-world laboratory results, overall and in transplant recipients; and (3) offer considerations for the interpretation of negative and low-positive serological results, particularly for high-risk individuals with increased vulnerability to SARS-CoV-2 infection and its consequences.

## Methods

### Evidence Review

A review of the scientific literature for the topic of SARS-CoV-2 serology and subsequent outcomes was conducted in March to July 2022 using the Scopus® (Elsevier) curated abstract and citation search engine, including the search terms COVID-19, SARS-CoV-2 antibody, immunosuppression, and COVID-19 outcomes. Abstracts of approximately 300 journal articles were initially retrieved and reviewed, of which 69 directly relevant to interpretation of SARS-CoV-2 spike-targeted antibody results to assess risk of infection and severe outcomes were critically reviewed for potential inclusion.

### Real-World Data Analyses of SARS-CoV-2 Spike-Targeted Antibody Testing Results

Real-world clinical laboratory data were evaluated to better understand spike-targeted antibody seroprevalence in the United States. We assessed deidentified data from patients who received SARS-CoV-2 spike protein-targeted, semiquantitative antibody testing at Quest Diagnostics and LabCorp from March 2021 through June 2022. The laboratory-based methods were the DiaSorin LIAISON® SARS-CoV-2 IgG TrimericS (Saluggia, Italy), reported as BAU/mL; Roche Diagnostics (Indianapolis, IN) Elecsys® Anti-SARS-CoV-2 S, reported as U/mL; and the Siemens Healthcare Diagnostics (Tarrytown, NY) ADVIA Centaur®/Atellica® SARS-CoV-2 IgG, reported as an index.^8^ Instrument values produced from the Roche and Siemens methods were converted to binding antibody units/mL (BAU/mL), based on the published conversion formulas.^9^

Antibody positivity rates in solid organ transplant recipients (an immunocompromised group at specific high risk of severe outcomes from COVID-19, given lifelong immunosuppression) were compared to the general population for the period when Omicron variants were dominant in the United States (January-June 2022). Individuals were categorized as transplant patients if they had an International Classification of Diseases, 10th Revision (ICD-10) code for transplant within 3 months prior to SARS-CoV-2 spike-targeted antibody testing (Table 1). Statistical analysis was performed using R version 3.4.0 (R Core Team, 2017). Seronegativity rates were compared applying chi-squared test and statistical significance was assumed for *P* values <.01.

**Table 1.** International Classification of Diseases, 10th Revision (ICD-10) Codes to Define Post-Transplant.

| **ICD-10 Code** | **Description** |
| --- | --- |
| T86.x | Complications of transplanted organs and tissue |
| Z482.x | Encounter for aftercare following organ transplant |
| D47Z1 | Post-transplant lymphoproliferative disorder (PTLD) |
| C802 | Malignant neoplasm associated with transplanted organ |

## Observations

### Risk Factors for Severe COVID-19 Illness

The following groups were identified as being more vulnerable to severe illness from COVID-19:^4^ (1) adults aged ≥65 (accounting for >80% of COVID-19-related deaths); (2) individuals with underlying medical conditions (ie, cancer, chronic kidney disease, chronic liver disease, cystic fibrosis, dementia or other neurological conditions, diabetes, disabilities, heart conditions, human immunodeficiency virus (HIV) infection, immunocompromised conditions or weakened immunity); and (3) individuals encountering barriers to healthcare access, including racial/ethnic minority groups and people with disabilities. Immunocompromising conditions, one of the main risk factors for vulnerability, can include active cancer treatment, organ transplant, immunosuppressive medications, stem cell transplant within the past 2 years, moderate to severe primary immunodeficiency (eg, DiGeorge syndrome, Wiskott-Aldrich syndrome), or advanced or untreated HIV infection.^10^

Multiple studies have documented increased risk of adverse outcomes in high-risk groups.^11-13^ Thus, appropriate primary vaccination and booster doses are particularly important for high-risk individuals and additional dosing may be beneficial if no antibody response is detected post-vaccination. The sections below discuss the impact of risk factors for increased vulnerability on vaccine response and effectiveness, as well as the potential role of serologic testing for assessing individual risk and informing the need for additional vaccine doses and/or other interventions.

### Vaccine Response and Effectiveness in High-Risk Individuals

Understanding the altered relationship between vaccine effectiveness and clinical outcomes in high-risk populations is essential for appropriate management and risk mitigation. Immunocompromised populations have been reported to have highly variable and suboptimal antibody responses to SARS-CoV-2 vaccination, with a correspondingly high rate of breakthrough infections (infection post-vaccination).^14^ Antibody responses may be especially variable for patients receiving immunosuppression medications for solid organ transplant.^15,16^ Transplant recipients have poorer vaccine response or delayed response.^17^ In turn, poor vaccine-induced antibody response has been associated with inferior clinical outcomes.^13,18^ Numerous studies have reported lower vaccine effectiveness in immunocompromised individuals compared to the general population.^5,19,20,21^ Shorter durability of NtAb responses was found among immunocompromised individuals (n=637) compared to healthy individuals (n=204), especially with the Beta and Delta variants.^22^ At 3 months post-vaccination, the percentages of patients with NtAb responses to the Delta variant fell to 88.9% among healthy controls and 38.8% and 16.1% for those with autoimmune diseases and solid organ transplants, respectively.^23^ Additionally, significantly higher mortality rates were reported among partially-vaccinated compared to fully-vaccinated patients with cancer.^23,24^

### Negative SARS-CoV-2 Serology Associated with Increased Risk of COVID-19 Infection

Studies have consistently demonstrated that negative antibody test results identify people at increased risk for subsequent infection.^25-28^ In a study of 19,000 unvaccinated participants, the rate of polymerase chain reaction (PCR) positivity at >90 days after an index antibody test result was 10 times higher for those who were initially antibody negative.^26^ Similarly, in a phase 3 efficacy trial of the mRNA-1273 vaccine, vaccine recipients were assessed for NtAb and binding antibodies as correlates for subsequent risk; all serological markers were inversely associated with COVID-19 risk and positively associated with vaccine efficacy.^29^

### Low-Positive SARS-CoV-2 Spike Binding Antibody Results and Risk of Breakthrough Infection

Beyond non-detectable (ie, negative) SARS-CoV-2 spike binding antibody levels, low-positive spike antibody levels may also signal increased vulnerability to SARS-CoV-2 infection.^30^ In a study of 1394 patients with hematological disorders, absence of detectable antibodies at 3 to 6 weeks post-vaccination was the only variable independently associated with breakthrough infections; incidence and severity of COVID-19 were significantly higher for patients with values <250 BAU/mL.^31^ In a study of 114 immunocompromised individuals, SARS-CoV-2 spike-targeted antibody levels were undetectable in 70% and <300 BAU/mL in 92% of hospitalized patients, at admission.^32^

A role for low-positive semiquantitative/quantitative SARS-CoV-2 serology results in patient monitoring has been recognized in guidelines from other countries, including the French Vaccine Strategy Guidance Council (COSV).^30^ For individuals with antibody levels >264 BAU/mL determined with a SARS-CoV-2 spike-targeted test (based on a study that included the Oxford Vaccine Trial Group^32^), protection from a booster vaccine dose is considered sufficient; additional serology testing is recommended every 3 months. A South Korean study found SARS-CoV-2 antibody levels are predictive of the clinical course of COVID-19 in vaccinated patients with Delta and Omicron variant infections. The study found enhanced viral clearance in vaccinated patients with higher antibody levels than in those with lower antibody levels.^33^

Barrière and colleagues suggested offering additional vaccine doses to non-responders (<40 BAU/mL) and those with intermediate SARS-CoV-2 antibody levels (40-100 or 40-260 BAU/mL).^34^ Individuals with higher antibody titers can follow general population guidelines or have successive SARS-CoV-2 antibody testing to monitor antibody levels. Wagner and colleagues evaluated vaccine effectiveness in patients with immunocompromised conditions, concluding that, “Immunomonitoring … seems advisable to identify vaccination failures and consequently establishing personalized vaccination schedules, including shorter booster intervals, and helps to improve vaccine effectiveness in all patients with secondary immunodeficiencies.”^35^

### Comparability of Quantitative and Semiquantitative SARS-CoV-2 Spike Binding Antibody Assays

For semiquantitative and quantitative SARS-CoV-2 spike-binding antibody assays to be used in clinical decision-making, understanding the comparability of results between different tests is important. Various FDA EUA assays provide different semiquantitative or quantitative unit scales. Among high-throughput semiquantitative SARS-CoV-2 spike binding antibody assays (eg, Abbott Laboratories, Beckman-Coulter, DiaSorin, Ortho-Clinical Diagnostics, Roche Diagnostics, and Siemens Healthineers),^36^ some have demonstrated strong qualitative performance in predicting surrogate virus neutralization seropositivity.^37,38^ The International Standard and International Reference Panel for anti-SARS-CoV-2 immunoglobulins, adopted by the World Health Organization Expert Committee on Biological Standardization,^39^ improves harmonization of assay results to an arbitrary BAU/mL. This harmonization reduced inter-laboratory variation and created a common language for reporting data. However, quantitative comparison between different commercial assays is contingent upon similar assay formats, viral protein targets, and antibody classes being detected by each method.^8^

Results from semiquantitative and quantitative SARS-CoV-2 spike-binding antibody tests correlate strongly with the presence of surrogate pseudovirus NtAb.^40^ Formal neutralization assay testing using spike variant peptides, as well as T-cell based immune function assays evaluating the adaptive immune response to SARS-CoV-2, may allow for more effective interpretation of immunity and/or future protection. However, these technically more sophisticated assays are not yet widely available and/or scalable to meet testing needs across the United States. Semiquantitative and quantitative FDA EUA serology assays that target spike-protein binding antibody have the advantage of providing some level of objective measurement of an adaptive immune response to both vaccination and natural infection. Khoury and colleagues demonstrated that, after normalization of SARS-CoV-2 antibody titers, “studies converge on a consistent [positive] relationship between antibody levels and protection from COVID-19.”^41^

### SARS-CoV-2 Spike-Targeted Antibody Results: Real-World Data

The CDC has estimated the national SARS-CoV-2 serology positivity rate by evaluating nucleocapsid-targeted serology testing performed by large commercial clinical laboratories on remnant specimens with all identifiers except state residence were removed.^42^ These data show that the seronegativity rate has moved from >90% in 2020 to between 66% to 71%% throughout 2021, and reached a low-point of at 42% in its last assessment period, January 27-February 26, 2022. Quest Diagnostics and LabCorp data for SARS-CoV-2 spike-targeted antibody testing, (2,970,722 results) (DiaSorin, Roche, and Siemens), showed a similar trend, with seronegativity of 40% in March/April 2021 and reaching <10% in March-June 2022 (Figure 1). Seronegativity rates have stabilized in recent months based on the net of vaccinations, new infections, and waning immunity.

**Figure 1.**
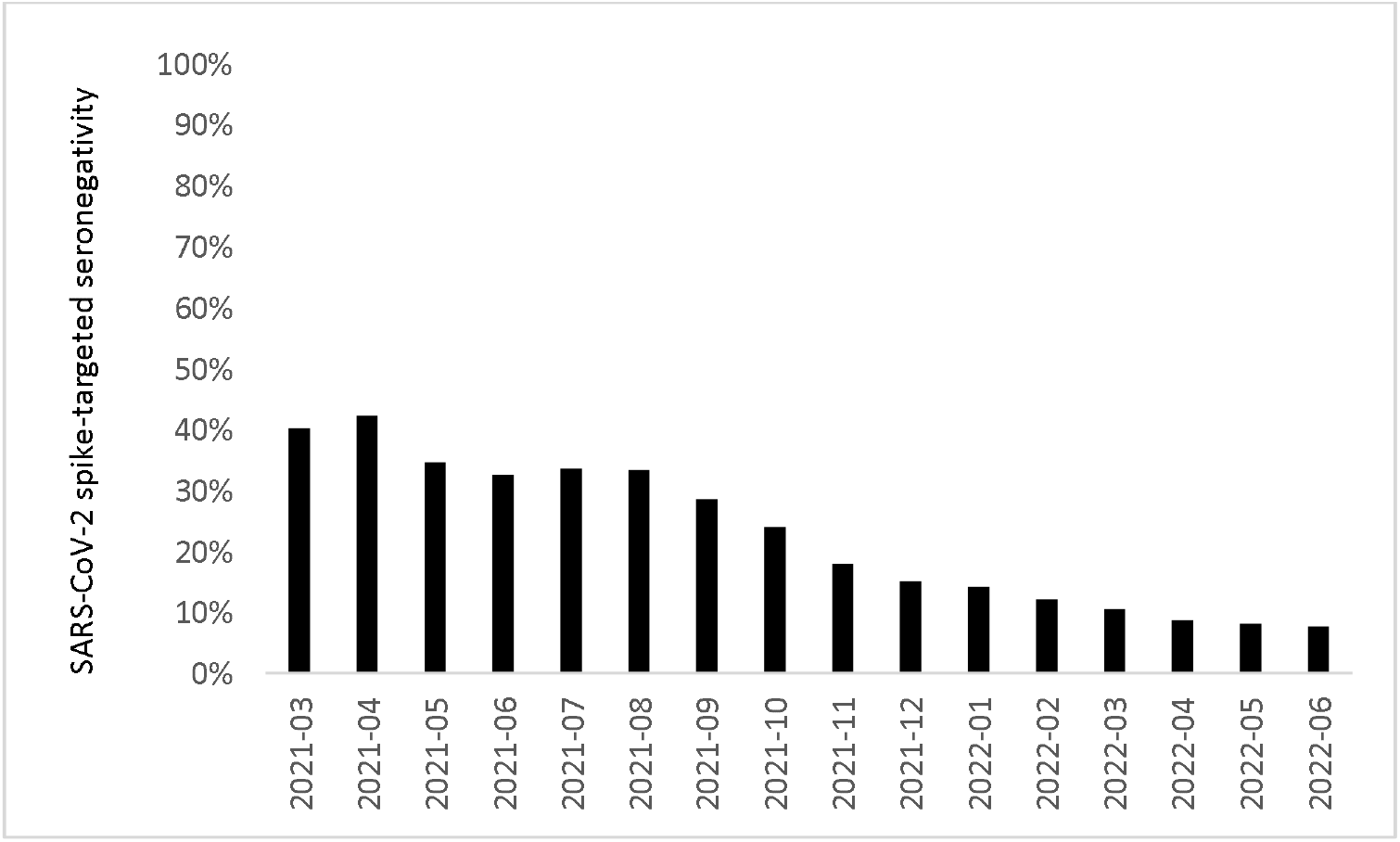
LabCorp and Quest Diagnostics Monthly SARS-CoV-2 Spike-Targeted Seronegativity Rate in the United States, March 2021 Through June 2022 (Based Upon 2,970,722 Tests)

A total of 4439 results were from patients tested at Quest Diagnostics with preceding ICD-10 codes for transplantation. The Roche test yielded antibody-negative results 42% more often in solid organ transplant recipients (7.7%; 115/1,492) than in the overall population (5.4%; 3724/68,481) (*P*<.01); the 20th percentile of results for the Roche test was also substantially lower in transplant recipients (98 BAU/mL) than the general population (250 BAU/mL) (Table 2). The Siemens test yielded negative results 76% more often for transplant recipients (21.1%; 627/2,967) than for others (12.0%; 43,828/364,000) (*P*<.01); the 20th percentile and median values of results were also substantially lower for transplant patients (45 and 854 BAU/mL, respectively) than for the general population (139 and 1664 BAU/mL, respectively).

**Table 2.**
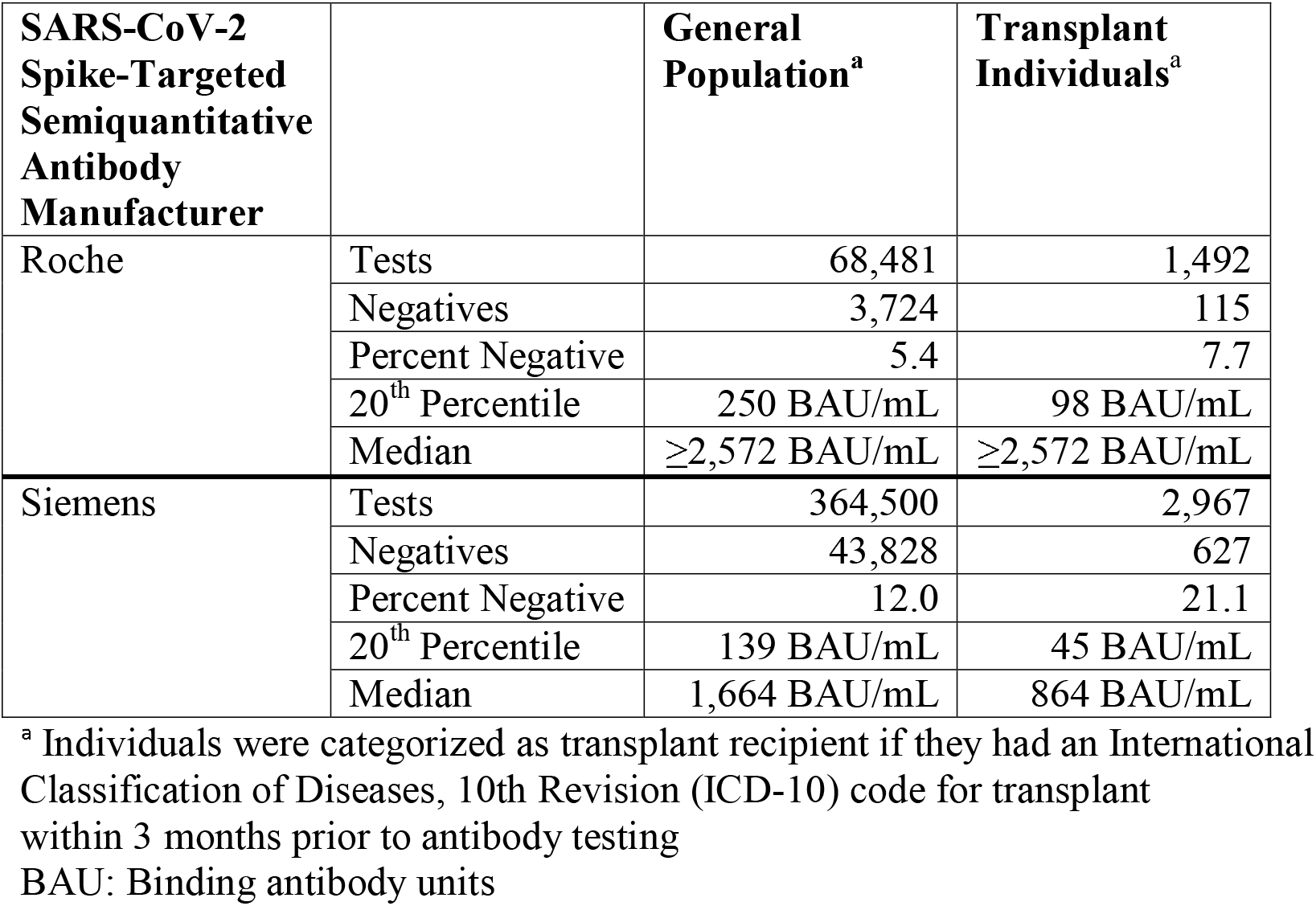
SARS-CoV-2 Spike-Targeted Antibody Test Results for Patients Categorized as Transplant or General Population, January 2022 Through June 2022.

## Discussion

Available evidence supports a role for assessment of SARS-CoV-2 spike-targeted antibody presence including quantification, in refining risk assessment for individuals at increased vulnerability to severe COVID-19 illness and death.^5^ Such assessment would provide physicians with actionable data that can be used with available clinical information to guide patient interventions.^43^ Health care providers seek detailed guidance from governmental health agencies on how to gauge the SARS-CoV-2 vulnerability of immunocompromised individuals and wider high-risk populations. Our experience is that many clinicians are developing their own SARS-CoV-2 serology result parameters to aid in decision-making, but without the benefit of expert guidelines on interpretation of specific antibody levels. We propose that, in the near term, commonly available SARS-CoV-2 spike-targeted antibody data and/or combinations with NtAb assays can be a valuable resource for clinicians and their patients who seek information regarding vulnerability to infection and indications for preventative therapeutics to avoid associated adverse outcomes. Tracking changes in semiquantitative SARS-CoV-2 spike-targeted antibody results may provide insights into the durability of antibody response. If semiquantitative or quantitative tests are used to evaluate vulnerability to SARS-CoV-2 infection, assay-specific vulnerability thresholds would be needed for each assay and may vary with the predominant circulating SARS-CoV-2 variants.

Establishing antibody response 2 to 4 weeks post-vaccination may be useful. Individuals at high risk because of being immunocompromised or other factors may benefit from frequent testing (every 2 to 3 months), given their potential for rapid declines in SARS-CoV-2 antibody response. Caution in this situation must be exercised because semiquantitative assays are nonlinear; changes are directional and not absolute.

As reviewed above, substantial literature supports the interpretation of negative SARS-CoV-2 spike-targeted antibody results to identify individuals with increased risk of SARS-CoV-2 infection and therefore potentially associated adverse outcomes.^19,20,21,22^ Interpretation of non-detectable SARS-CoV-2 spike-targeted antibody to indicate increased risk represents a clinically actionable approach to serology testing, one that aligns with established recommendations for vaccination and behavioral interventions to reduce the spread and impact of infection. This approach may also encourage greater adoption of measures to protect individuals (eg, booster vaccinations and personal protective measures). In support of this concept, a recent expert position statement strongly recommended that patients with cancer be included in observational serological monitoring studies or clinical trials dedicated to severe immunocompromised patients without evidence of a serological response following additional vaccine doses.^34^ In a study of patients on hemodialysis, the authors strongly recommended use of SARS-CoV-2 serological monitoring after vaccination to determine booster timing, especially for patients with malnutrition or on immunosuppressive therapy.^44^

Patients with negative results on SARS-CoV-2 spike-targeted antibody tests may also benefit from supplemental tests to better understand their vulnerability to SARS-CoV-2 infection and adverse outcomes. Alternative assays that measure antibody neutralization of novel spike protein(s) or cellular-based adaptive immunity assays are being studied but are not yet widely available.^45^

The evidence discussed above suggests that a negative SARS-CoV-2 spike-targeted serology result identifies individuals, among those with immunocompromised or high-risk conditions, as having higher risk for SARS-CoV-2 infection. With this approach, a negative result on a laboratory-based FDA EUA SARS-CoV-2 spike-targeted antibody test (≥4 weeks after full vaccination or in an unvaccinated individual) identifies individuals at highest risk of SARS-CoV-2 infection and associated adverse clinical outcomes. Piñana and colleagues showed that, among patients with hematological diseases, SARS-CoV-2 antibody levels 3 to 6 weeks post-2 dose vaccination were associated with protection against both breakthrough infections and severe disease from Omicron variants.^31^ Our estimates based on commercial laboratory seroprevalence studies and blood donor surveys^46^ suggest that, overall, approximately 5% to 25% of the US population have SARS-CoV-2 spike-targeted antibody levels below positivity thresholds on commonly used tests, with disproportionately high rates in immunocompromised populations.^47^ The real-world SARS-CoV-2 spike-targeted antibody data from 2,970,722 patient results presented (Figure 1) demonstrate that seronegativity was just <10% during April through June 2022. While most people demonstrate an immune response, some groups, particularly those at high risk because of immunosuppression or other conditions, may not, even with prior vaccination or exposure. Solid organ transplant recipients were more likely than the general population to have negative SARS-CoV-2 serology results and had generally lower semiquantitative levels. Note that the presented data were collected primarily from clinician-ordered serology tests, most likely after patient vaccination. Therefore, seronegativity is likely significantly more common among the unvaccinated, non-infected general population.

Differences in positivity rates among the employed commercial serology methods in this report may reflect differences in epitopes, assay formats, criteria for defining negativity, and possibly populations tested. However, our observations among transplant patients are consistent with those of previous studies showing that immunocompromised patients are more likely to have non-detectable SARS-CoV-2 spike-targeted antibodies and lower semiquantitative antibody responses.^20,21,22,23^

Low-positive SARS-CoV-2 spike-targeted antibody results (eg, in the 240-264 BAU/mL range^40,41,42^) may also provide clinical insight. The precise level defining a low-positive threshold varies across studies but cluster around 260 BAU/mL; this level may hold potential as a threshold to identify additional individuals who are antibody positive but still at higher risk of infection and subsequent potential adverse outcomes. Consideration of individuals with low-positive SARS-CoV-2 spike-targeted antibody levels would increase the percentage of the population classified as having greater vulnerability to SARS-CoV-2 infection and potential adverse outcomes, and potentially aid in prioritizing them for enhanced measures to prevent COVID-19 or mitigate its clinical course.

Further data are needed to more precisely delineate the risk of adverse clinical outcomes, such as hospitalization and death, associated with negative or low SARS-CoV-2 spike-targeted antibody results. One additional challenge for semiquantitative and quantitative testing in assessing vulnerability to severe COVID-19 illness is how new variants might affect thresholds for what is considered a low SARS-CoV-2 antibody level (relative to spike protein(s) neutralization potential). Since new SARS-CoV-2 variants appear to possess increased immune escape capabilities, this current approach would likely remain clinically useful, especially if SARS-CoV-2 spike-targeted antibody levels are reflective of potent neutralization levels of antibody needed to contain viral replication. Though, a recent study suggests immune escape of new variants must be considered.^48^

## Conclusions

Individuals with a negative SARS-CoV-2 spike-targeted antibody test result are at the highest risk for SARS-CoV-2 infection with potential adverse outcomes. SARS-CoV-2 semiquantitative or quantitative SARS-CoV-2 spike-targeted antibody testing provides a potentially important objective guidepost to evaluate the immune response and assist in the care of high-risk groups. This information will be most useful when evaluated by a clinician considering a patient’s overall clinical risk for adverse outcomes. The role of negative, and possibly low-positive, results deserve further study for their potential to identify individuals at increased risk for severe COVID-19 illness. Nevertheless, we await additional data to further refine the association of specific SARS-CoV-2 spike-targeted antibody levels and/or their neutralization potential with the degree of subsequent infection/disease risk.

## Data Availability

All data produced in the present work are contained in the manuscript

## Data Availability

All data produced in the present work are contained in the manuscript

## Author Contributions

Kaufman and Meyer III had full access to all the data in the study and take responsibility for the integrity of the data and the accuracy of the data analysis.

*Concept and design:* Kaufman, Meyer III, Wohlgemuth, Clarke

*Acquisition, analysis, or interpretation of data:* Kaufman, Meyer III, Clarke, Radcliff, Rank, Freeman, Eisenberg, Gillim, Morice, Briscoe, Kenyon, Perlin, Wohlgemuth

*Drafting of the manuscript:* Kaufman, Meyer III, Wohlgemuth

*Critical revision of the manuscript for important intellectual content:* Kaufman, Meyer III, Clarke, Radcliff, Rank, Freeman, Eisenberg, Gillim, Morice, Briscoe, Kenyon, Perlin, Wohlgemuth

*Statistical analysis:* Kaufman

*Administrative, technical, or material support:* Radcliff

*Supervision:* Wohlgemuth

